# Changes in local immune status in tonsillopharyngitis under pathogenic therapy with the BNO 1030 extract: a randomized, comparative, parallel-group, single-center study

**DOI:** 10.1101/2023.07.28.23293249

**Authors:** Elena L. Savlevich, Vladimir S. Kozlov, Andrey M. Gaponov, Andrey N. Gerasimov, Petr V. Markus, Elena V. Yakushenko, Natalia E. Doroshchenko, Ivan G. Kozlov

## Abstract

**Aim of study:** The present study aimed to evaluate anti-inflammatory and immunomodulating properties of BNO 1030 (Tonsilgon® N) and its direct effect on the indicators of the local immunity of oropharyngeal mucosa in patients with acute tonsillopharyngitis (TP) or exacerbation of chronic TP without evident systemic inflammatory syndrome.

**Materials and methods:** A total of 60 adult patients with acute TP or exacerbation of chronic TP without severe systemic inflammatory syndrome were randomly divided into 2 groups: Group 1 L 30 patients took BNO 1030 (Tonsilgon® N), Group 2 L 30 patients took sage tablets according to the summary of product characteristics during 7 days. During 3 visits (day 1, day 3, day 7) symptoms and oropharyngeal mucosa condition were evaluated using a 10-point visual analogue scale (VAS). Local immunity parameters of oropharyngeal mucosa (cytokines: IL-1β, IL-6, IL-8, IL-10, IL-17, TNF-α, and lysozyme, lactoferrin, sIgA) were determined by ELISA and by real time polymerase chain reaction.

**Results:** Reduction of the main symptoms was significantly faster under BNO 1030 therapy than under sage therapy. In BNO 1030 group reduction of clinical symptoms correlated with the onset of action and the local immunological parameters. During BNO 1030 treatment IL-1β, IL-6, and IL-8 mRNA levels decreased below the levels in healthy controls, while, the immune factors lysozyme, lactoferrin and sIgA increased. Therapy with sage tablets did not affect local immunity parameters.

**Conclusion:** Both treatment regimens resulted in elimination of clinical signs and mucosal pharyngeal barrier regeneration. In contrast to the sage tablets, BNO 1030 can also affect local mucosal immunity via regulating the balance of pro- and anti-inflammatory factors.

**Study highlights:**

- This study can answer the question regarding influence on main local immunity parameters of subjects with tonsillitis
- The one of main task of this study is to evaluate what local immune parameters of oral cavity are most changeable
- Tonsilgon N usage is not decrease main local immunity parameters such as IL-1β, IL-8, lysozyme and lactoferrin
- This study evaluated the functional condition of mucosal immunity of oral cavity in subjects with tonsillopharingitis

## Introduction

Acute inflammatory diseases of ear nose throat (ENT) organs are one of the actual problems of modern public health care due to the leading position in treatment demand, temporary disability and number of used medicines in all age categories. Almost 100% of patients with acute respiratory diseases complain of painful swallowing, burning, throat irritation, dryness and presence of foreign body in oropharynx, which are clinical signs of acute tonsillopharyngitis (TP) or exacerbation of TP [1]. It is well known that inflammatory diseases of the oropharynx in 70-85% have a viral etiology with a predominance of rhinoviruses, adenoviruses, coronaviruses, respiratory syncytial viruses and influenza viruses. A separate group of herpes viruses includes Epstein-Barr virus, in which TP is accompanied by generalized lymphadenopathy and hepatosplenomegaly [2]. Moreover, as an isolated agent or as a co-infection with respiratory viruses, bacterial agents, including β*-hemolytic Streptococcus* group A, *Staphylococcus aureus, Streptococcus pneumoniae, Mycoplasma pneumoniae, Chlamydia pneumoniae* and *Arcanobacterium haemolyticum* can be a trigger for TP [3].

The initial link in the pathogenesis of TP of any etiology is an immunoinflammatory process at the site of entry gate of infection, and the dominant symptom causing significant adverse effect on quality of life is painful swallowing, which can be manifested severely due to specific features of innervation of this anatomical region [4]. When inflammation progresses, hypersensitivity of pain receptors in the posterior pharyngeal mucosa develops, which increases pain sensitivity and makes it difficult to eat and even to swallow minimal volumes of fluid. Inflammation, however, is a nonspecific reaction of the body that develops according to standard pathways. The changes that occur in tissues during influenza have been studied most extensively in the past. Toxins, products of tissue breakdown by cytolysis or apoptosis of cells due to massive release of mature virions and other triggers induce epithelial cells and macrophages to synthesize chemokines, monocyte chemotactic protein (МСР)-1, MCP-3, regulated upon activation, normal T-cell expressed and secreted (RANTES), and interleukin (IL)-8. Chemokines in turn induce migration of immunological active cells into the inflammation focus, directly initiating and participating in local inflammatory process – neutrophils, eosinophils, macrophages, lymphocytes. This leads to the production and release of biologically active substances that intensify alteration phenomena, destruction, stimulate synthesis of acute-phase proteins and oxidative stress. This process is also known as early cytokine response phenomenon, which is a nonspecific body response resulting from the interaction of infectious agents with macrophages and other effector cells. This results in activation of natural killer cells (NK), redistribution and differentiation of certain lymphocyte subpopulations, gene expression followed by synthesis of interferon (IFN)-α/β/γ, IL-1, IL-6, IL-8, IL-12, IL-18, and tumour necrosis factor (TNF)-α proteins [5]. Subsequently, excessive synthesis of proinflammatory cytokines manifests their systemic effects in the form of a general intoxication syndrome manifested by elevated body temperature, chills, headache, heaviness in the head, malaise, fatigue, increased sweating, loss of appetite, nausea. However, in TP without a pronounced systemic inflammatory syndrome, such studies are not presented.

Modern medicine has an abundance of pharmacological agents used for treatment of inflammatory processes in oropharynx, while at the same time, in the case of an acute pharyngitis on the out-patient stage, antibiotics are prescribed in 65-85% of cases (of which parenterally - more than 40%). In hospitals 98% of TP cases (parenterally - 90%) are treated with antibiotics, although, because of its viral etiology, this is neither justified nor expedient [6]. Moreover, the incidence of unnecessary antibiotic prescribing in children in the first year of life is 66.7% [7]. Besides the risk of antibiotic resistance development, patients show immunosuppressive effects associated with the release of bacterial toxins during the destruction of bacteria. Additionally, there is a decrease in the synthesis of local IFN-α/β, nonspecific protective factors, including lysozyme, secretory immunoglobulin A (sIgA), decreased T-lymphocyte numbers and IgG levels. The second problem in the treatment of TP is use of immunomodulatory agents, which are registered in the Russian Federation in excess of a hundred [8].

However, human resistance to various respiratory infectious agents is a multifactorial phenomenon. Factors of innate and adaptive immunity and respiratory epithelia are closely interrelated and complement each other, representing only individual links of the protective mechanism, which is the anti-infective immunity. The leading role is given by the first line defense of the mucosal immunity of the oropharynx, represented by antimicrobial peptides like lysozyme and lactoferrin as well as sIgA and interferons. These factors are involved during the complete process of inflammation [9]. They form a protective barrier to viruses much earlier than specific immune defenses by stimulating cellular resistance, rendering cells unfit for viral multiplication, and the course and outcome of disease depend on the speed with which the interferon system is incorporated into the antiviral defense [10]. In acute nasopharyngitis with systemic inflammatory syndrome in the first 2 days of the disease no deviations from the normal range of parameters of systemic humoral and cellular immunity can be revealed. Only a decreased relative number of cytotoxic T-lymphocytes (CD3+CD8+), stimulated nitroblue tetrazolium test (NST-test) can be noted, indicating phagocytic activity of granulocytes and increased phagocytic activity of monocytes. At the same time in nasal mucus a decrease of indicators of the local immune response such as IFN-α and sIgA content can be determined [11]. Therefore, studying of indexes of the local immune status of the oropharyngeal inflammatory pathology gives the most information on local immune defense mechanisms, anyway this process has not been studied and discussed enough.

The history of phytotherapy application in the treatment of oropharyngeal diseases has a centuries-long duration due to which in modern conditions the spectrum of plants having the highest efficiency in the elimination of certain clinical manifestations is already highlighted. Due to the fashionable tendencies to follow a healthy lifestyle at the present time in many countries, there is an active search for the possibility of using medicinal plants as an alternative method or as an “add-on” approach for the treatment of many pathological conditions. Nowadays the development of science and the pharmacological industry make it easily achievable to create modern and innovative technologies for standardization of herbal medicinal raw materials including quality processing of the initial material in the production process of preparations. The main problem of phytotherapy remains the difficulty of determining the mechanisms of action of a particular drug with proven clinical efficacy in specific indications.

A similar situation is observed with the herbal medicinal product BNO 1030 (Tonsilgon^®^ N), drops for oral administration (Bionorica SE, Germany), which contains water-alcoholic extracts of marshmallow root (*Althaea officinalis*), Chamomile flowers (*Chamomilla recutita*), horsetail herb (*Equisetum arvense*), walnut leaves (*Juglandis regia*), yarrow herb (*Achillea millefolium*), oak bark (*Quercus robur*), dandelion herb (*Taraxacum officinale*). Its efficacy in treatment of inflammatory phenomena of the oropharynx and acute respiratory viral infections has been repeatedly shown in many clinical studies, but the mechanism of action of this drug in these diseases to date remains poorly understood.

**The aim of this work** was to study anti-inflammatory and possible immunomodulating properties of BNO 1030 and its direct effect on the indicators of the local immunity of oropharyngeal mucosa in patients with acute TP or exacerbation of chronic TP without evident systemic inflammatory syndrome.

## Materials and methods

### Trial Design

An interventional, investigator-blinded, laboratory-level, randomized, parallel-group comparative study (ISRCTN registry #ISRCTN80067058, https://doi.org/10.1186/ISRCTN80067058) was conducted from 01-Nov-2019 to 31-Dec-2020. The subjects’ recruitment has been lasted since 01-Jan-2020 till 23-Dec-2020. The clinical study was carried out in compliance with the WMA Declaration of Helsinki - Ethical Principles for Medical Research Involving Human Subjects, 2020, the Protocol of the Council of Europe Convention on Human Rights and Biomedicine (1999), the Federal Law “On the Fundamentals of Public Health Protection in Russia” No. 323 (05/26/2021) and Federal Law No. 61 about Drugs Circulation. The study was approved by the Local Ethics Committee.

The study was conducted and all subjects included at the clinical base of Federal State Budgetary Institution Policlinic #3 (31 Grokholsky lane, Moscow 129090; Ph.: +7 495 9826571). The study сonducting was approved on 27/12/2019, by Ethics Committee at Federal State Budgetary Institution Policlinic #3 (31 Grokholsky lane, Moscow 129090; +7 495 9826571), ref: 1-12-2019. Before inclusion in the study and the first study procedure conducting, all patients were given an written informed consent form for review and careful reading, which was signed by them in two copies after the doctor’s answers to all their questions about the study and procedures details. Only after signing informed consent forms, the subjects were randomized using simple randomization list and divided into 2 groups. Group 1 consisted of 30 patients (10 men, 20 female, mean age 41.5 with standard error of mean 1.93 years) took BNO 1030 according to the summary of product characteristics (SmPC), i.e. 25 drops orally every 2 hours during the first 3 days of treatment, then 25 drops 3 times a day up to and including day 7 as an alcohol solution for oral administration 30 minutes before or after a meal. Group 2 - 30 patients (11 men, 19 women, age 40.43 with standard error of mean 2.08 years) took sage tablets (containing sage dry extract (25 mg) and essential sage oil (2.4 mg) per tablet; Naturprodukt Europa b.v.; The Netherlands), 1 tablet for oral resorption every 2 hours during the first 3 days of treatment, then 1 tablet 3 times a day until and including day 7 (Figure 1). As acute tonsillopharyngitis is selflimited disease, most popular remedy officially recommended by guidelines in many countries are non-steroid anti-inflammatory drugs, that have at the same time, a fairly large number of side effects. Thus one of the study aims was evaluation of effectiveness and selection of most safe treatment, that is why herbal medicinal products officially used for acute tonsillopharyngitis treatment were chosen as study drugs.

**Figure 1:**
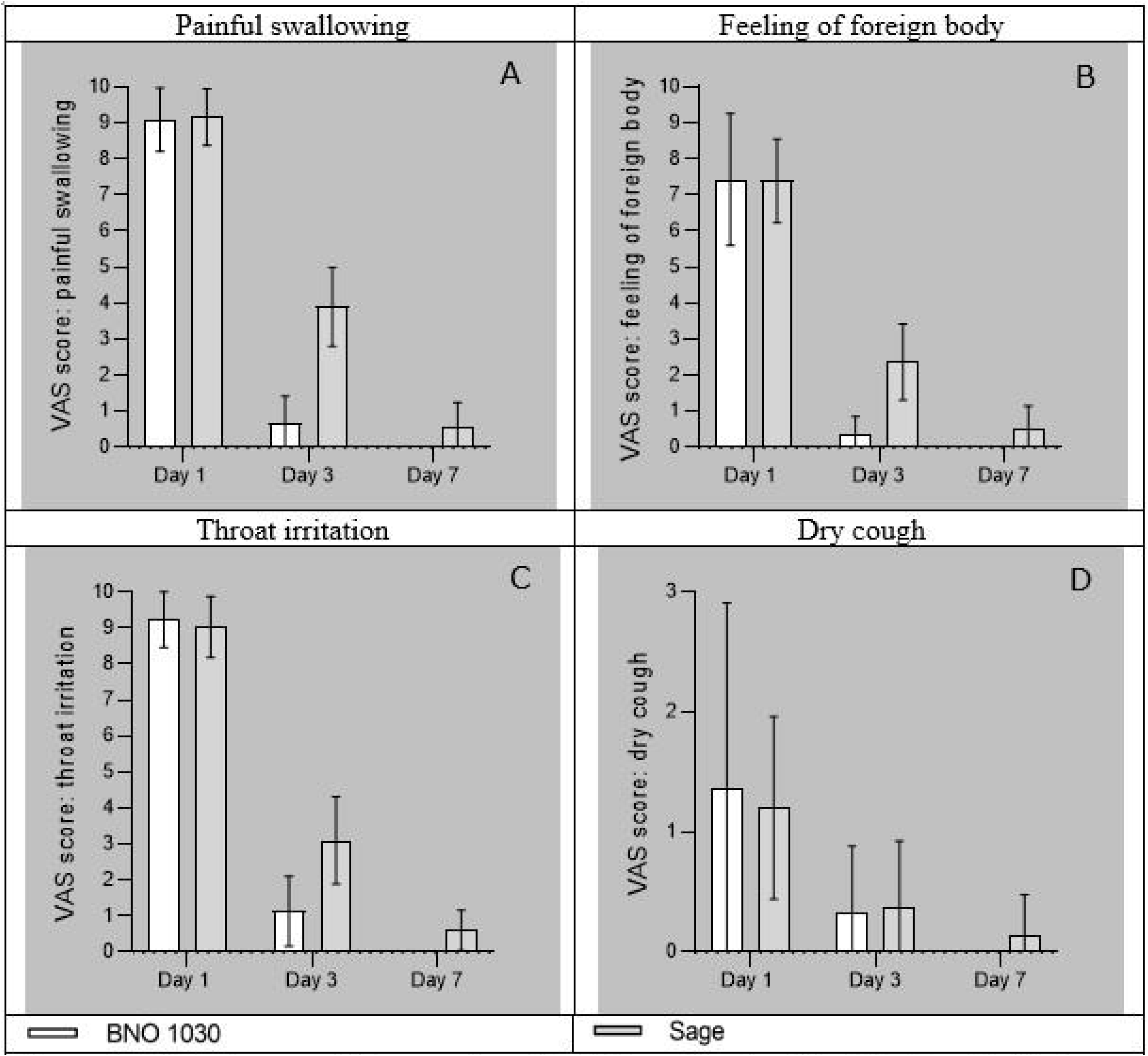
Clinical scores of the symptoms “painful swallowing” (A), “feeling of a foreign body in the oropharynx”(B), “throat irritation”(C), and “dry cough”(D) during the study according to a 10 point VAS assessment by the patients. Data are presented as means ± SD of N=30 patients per group. White bars represent parameters under BNO 1030 treatment, grey bars that under sage lozenges treatment

Since the reference range of oropharynx mucous membrane indexes of local immunity is absent, a control group was taken into the study. Control group L 30 persons (12 men,18 women, aged 40.17 with standard error of mean 1.81 years); they had no inflammatory manifestations of the respiratory tract at the time of material collection and within 6 months before this time, did not differ from the main group of patients by sex and age, and had no non-inclusion criteria.

### Participants

#### Inclusion Criteria

After signing informed consent, 60 adult patients with acute TP or exacerbation of chronic TP without severe systemic inflammatory syndrome (body temperature measured at axillary area ≤38.0°C), who had a maximum of 24 hours from the first symptoms of TP to contact a physician, were included in the study.

#### Exclusion criteria

Exclusion criteria were any signs of *Streptococcal* or fungal TP, diphtheria, rhinitis, sinusitis, otitis media, eustachianitis, laryngitis, tracheitis, bronchitis, taking antibiotics, local therapy for oropharyngeal diseases for < 48 hours before inclusion, systemic, inhaled or nasal glucocorticoids within 30 days prior to study inclusion, any co-morbidities requiring immunomodulators, pregnancy, lactation.

### Outcome measures

To determine the intensity of clinical manifestations and changes of the oropharyngeal mucosa, a pharyngoscopy was performed by an otorhinolaryngologist on visit 1on day 1, on visit 2 on day 3 and on visit 3 on day 7 of the disease. Symptoms and oropharyngeal mucosa condition were evaluated each time using a 10-point visual analogue scale (VAS) from 0 (no signs) to 10. The intensity of TP symptoms – painful swallowing, throat irritation, feeling of foreign body in oropharynx, dry cough, intoxication syndrome in form of fatigue, headache, body temperature increase - was assessed. Among objective signs the degree of hyperemia of the mucous membrane of the back wall of the pharynx and palatine tonsils was studied. For the immunologic part of the study, swabss were taken with a medical probe 3 times, on the days of patients’ visits, from the mucosa of the posterior pharyngeal wall and the palatine tonsils. The material was transferred into a sterile plastic microtube containing 1 ml of phosphate-salt buffer, pH=7.4, followed by centrifugation at 10’000 g (Eppendorf 5430R, Eppendorf AG, Germany) for 10 minutes. The supernatant and precipitate were collected separately and frozen at -80 °C for determination of proteins and mRNA expression of humoral factors by enzyme-linked immunoassay (ELISA) proteins and PCR-PCR (mRNA), respectively. The precipitate was lysed with RNA extraction reagent (CJSC Eurogen, Russia). before freezing. Levels of cytokines (IL-1, IL-6, IL-8, IL-10, IL-17, TNF-α) (Invitrogen, USA) and effector humoral immune factors (lysozyme, lactoferrin, sIgA) in supernatants were determined by ELISA according to the manufacturer’s protocol (CUSABIO Technology LLC using BMG Labtech, ClarioStar instrument (BMG Labtech AG, Germany). For standardisation, patient results were normalised by the determination of mucin content in each individual sample (LSBio Inc, USA). All protein data are presented in pg/ml.

Cytokine gene expression analysis (IL-1, IL-6, IL-8, IL-10, IL-17, TNF-α) was performed by RT-PCR using Eurogen kits (CJSC Eurogen, Russia) and primers according to Table 1 (Primer and probe sequences for the detection of cytokine gene expression levels by RT-PCR mRNA PCR primer pairs).

**Table 1:**
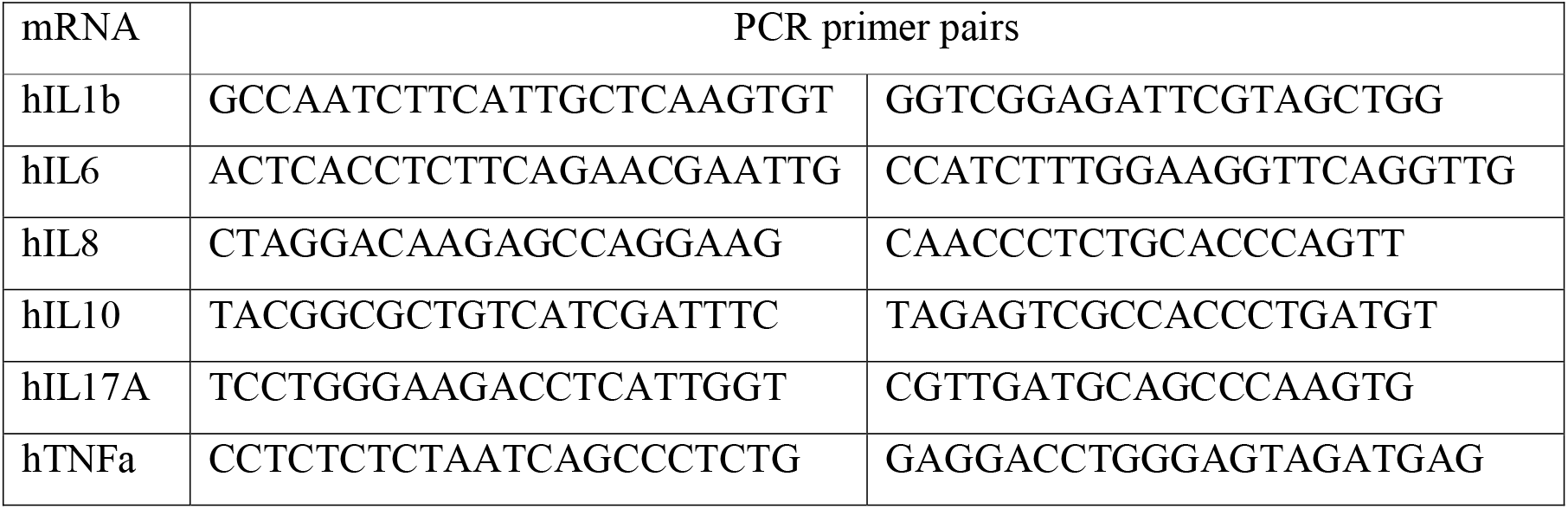
Primer and probe sequences for the detection of cytokine gene expression levels by RT-PCR mRNA PCR primer pairs.

All data are presented in conventional units. Transformation of mRNA into cDNA was performed on an IQ5 amplifier (BioRad, USA) with a set of laboratory reagents for total RNA isolation (QIAGEN, Germany), OT-1 reagent kit for reverse transcription (OOO NPF Sintol, Russia) and a set of reagents for RT-PCR (OOO NPF Sintol, Russia). All data of gene expression are presented as relative gene expression in units relative to the control group (Relative Quantity - RQ). The RQ values for the control group were taken as 1 and relative to it, the gene expression in other experimental groups was calculated.

No major study protocol deviations were registered during the study.

### Statistical Methods

Mathematical processing of the data was performed using IBM SPSS Statistics 23.0. The significance of the difference in frequencies between the compared groups was determined using the chi-square test (for the two groups, Fisher’s exact solution). Sufficiently large size of compared groups and small values of asymmetry coefficients of kurtosis allowed using methods of parametric statistics [12]. For numerical data and data in scores, their characteristic values are presented as mean and standard deviation (±) and median with 25^th^ and 75^th^ percentiles. Analysis of variance and the nonparametric Mann-Whitney test were used to compare the distributions of the two groups. A nonparametric Kruskal-Wallis test was used to compare the distributions of the numeric values between the three groups.

## Results

The differences in the proportions of males and females were not statistically significant when comparing the three groups: controls, BNO 1030 treatment, and sage treatment (p=0.866). Differences in the age distribution between the three groups were statistically insignificant, with p=0.890. The age distributions for males and females were also statistically insignificant (union of all three groups, p=0.184).

### Assessment of the evolution of clinical symptoms of TP

Assessment of clinical signs of TP on the 1^st^ day of treatment using a 10-point VAS scale (painful swallowing, feeling of dryness/smarting and feeling of foreign body in oropharynx, dry cough, fatigue, headache, increased body temperature, hyperemia, and throat irritation) revealed no statistically significant differences (p>0.05) between the two treatment groups. Further follow-up examinations were carried out on days 3 and day 7 to assess the efficiency of the treatment. Severities of the different symptoms by day are summarised in Table 2 (Dynamics of clinical manifestations of tonsillopharyngitis on the VAS scale and temperature in the 2 groups of patients with TP) and depicted in Figures 2 (Clinical scores of the symptoms “painful swallowing” (A), “feeling of a foreign body in the oropharynx”(B), “throat irritation”(C), and “dry cough”(D) during the study according to a 10 point VAS assessment by the patients) and 2 (Clinical scores of the symptoms “fatigue”(A), and “headache”(B) during the study according to a 10 point VAS assessment by the patients, as well as scores for “hyperemia of the mucous membrane”(C) (investigator’s assessment), and body temperature [°C] (D)).

**Figure 2:**
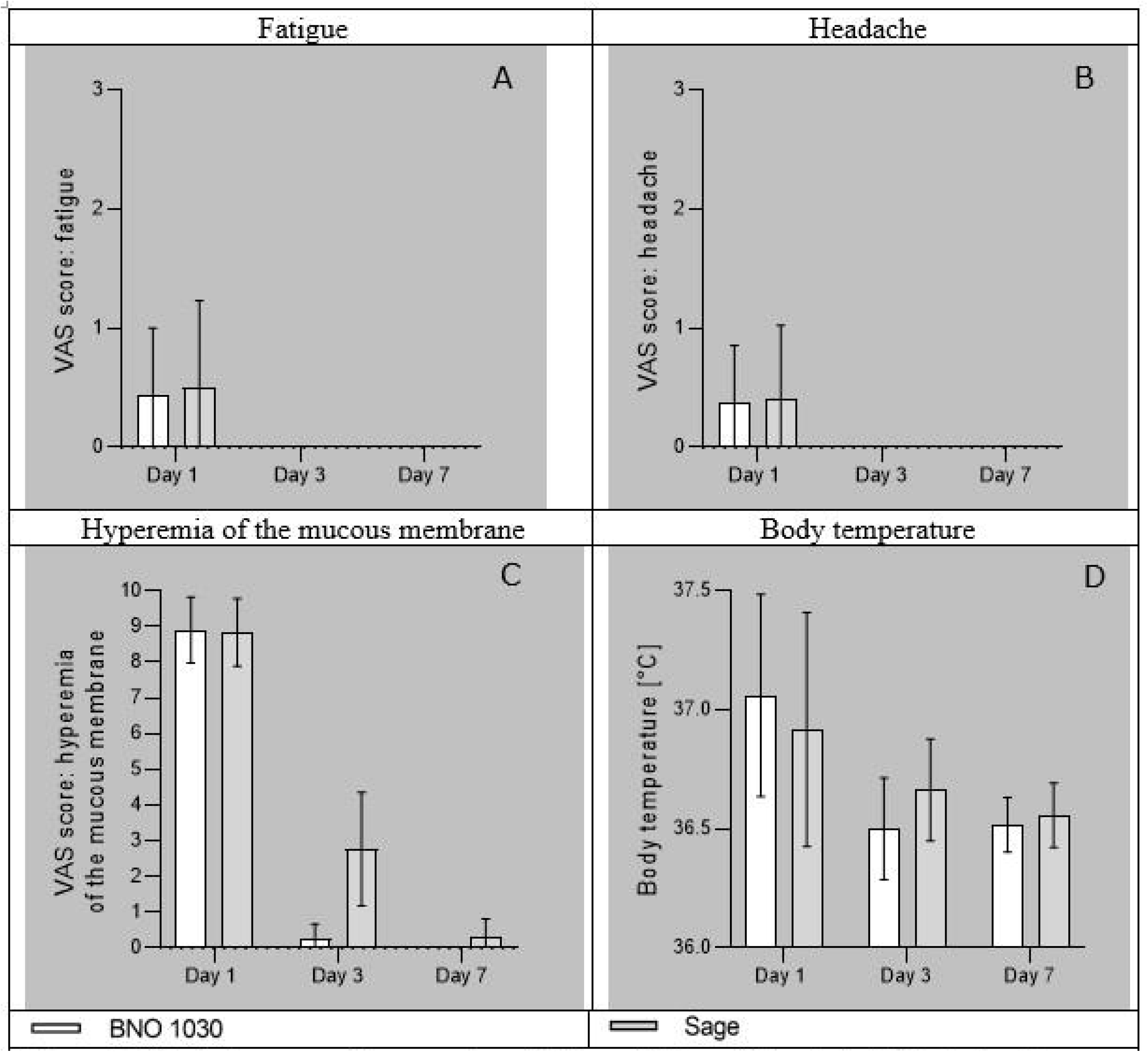
Clinical scores of the symptoms “fatigue”(A), and “headache”(B) during the study according to a 10 point VAS assessment by the patients, as well as scores for “hyperemia of the mucous membrane”(C) (investigator’s assessment), and body temperature [°C] (D). Data are presented as means ± SD of N=30 patients per group. White bars represent parameters under BNO 1030 treatment, grey bars that under sage lozenges treatment

**Table 2:**
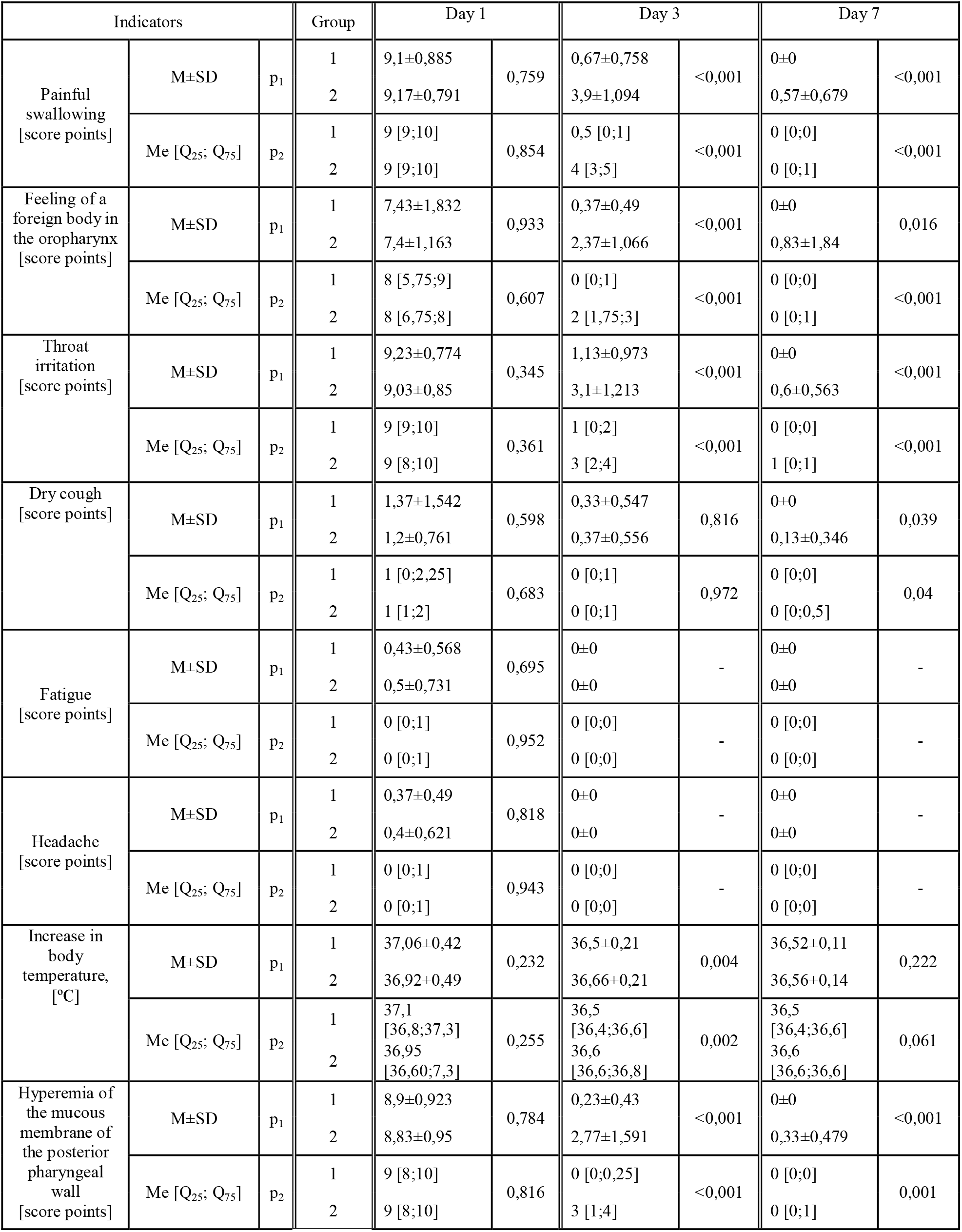
Dynamics of clinical manifestations of tonsillopharyngitis on the VAS scale and temperature in the 2 groups of patients with TP Data are presented as mean±standard deviation (Mean ± SD) and as Median (Me), [25th-75th percentile]. p < 0.05 indicates statistical significance. The significance of differences in the mean (p1), the significance of differences in the distributions (p2).

As can be deduced from Table 2 (Dynamics of clinical manifestations of tonsillopharyngitis on the VAS scale and temperature in the 2 groups of patients with TP), both treatment groups had a slight increase in temperature and significant severity of clinical manifestations of TP at the beginning of treatment. Further severity of clinical manifestations decreased and by day 7 in most cases almost completely regressed. On the third and the seventh day of treatment the progress in the BNO 1030 group was in most cases statistically more pronounced and significant.

As mentioned above, the main symptom delivering the most discomfort in TP is painful swallowing. In our study, 100% of patients with TP had this complaint. On the background of treatment there was a regress of this symptom in patients in both groups with a significant difference at the 2^nd^ and 3^rd^ visits: in the BNO 1030 group from a median score of 9 [7;10] on day 1 to 0.5 [0;1] on day 3 of the disease and absence of this complaint in 100% of cases on day 7. The sage group also showed regress of painful swallowing from median score points 9 [7;10] before treatment to 3 [2;4] points on day 3 and 0 [0;1] points on day 7; this symptom persisted in 46.7% of the group (Figure 2A, Clinical scores of the symptoms “painful swallowing, during the study according to a 10 point VAS assessment by the patients). A comparison of the severity for the parameter “feeling of a foreign body in the oropharynx”, also occurring in 100% of cases on the 1^st^ day of treatment, revealed a significant difference in both groups on days 3 and 7 of the treatment. The BNO 1030 group scored 8 [4;10] at the 1^st^ visit, 0 [0;1] at the 2^nd^ visit and was also asymptomatic on the 7^th^ day. In the sage group this symptom had a median score of 8 [5;9] points at visit 1, 2 [1.75;3] points at visit 2, and 0 [0;1] points at visit 3; this symptom persisted in 36.7% of the group (Figure 2B, Clinical scores of the symptoms “feeling of a foreign body in the oropharynx” during the study according to a 10 point VAS assessment by the patients). The symptom of throat irritation (sensation of dryness, burning sensation in the oropharynx) was noted in 100% of cases in both groups on the 1^st^ day of treatment. Comparison of symptom value rates also revealed a statistically significant difference at the 2^nd^ and 3^rd^ visit between the groups. On day 1 of referral median symptom severity was 9 [9;10] *vs.* 9 [8;10], on day 3 it was 1 [0;2] *vs.* 3 [2;4] in the BNO 1030 and sage group, respectively. On day 7 it was absent in the BNO 1030 and present in 56.7% (0[0;1] points) in the sage group (Figure 2C, Clinical scores of the symptoms “throat irritation”(C) during the study according to a 10 point VAS assessment by the patients).

A dry cough due to cough receptor irritation of the posterior pharyngeal wall was unexpressed on the day of presentation and occurred in 40% of cases. On the first 2 visits there was no significant difference in the decrease of symptom severity between the groups: on the first treatment day its median severities were 1 [0;2.25] score point *vs.* 1 [1;2] point in both groups. On the 2^nd^ visit there were 0 [0;1] median score points in both groups, on the 3^rd^ visit there were no symptoms anymore in the BNO 1030 group, while in the sage group it was observed in 13.3% of patients (median severity score 0 [0;0.1] points) (Figure 2D, Clinical scores of the symptoms “dry cough”(D) during the study according to a 10 point VAS assessment by the patients).

The manifestations of systemic inflammatory response also showed an active decline in both groups. There was no statistically significant difference in the reduction of fatigue and headache between the groups during all days of observation. On day 1 of observation the intensity of fatigue and headache was graded with median scores of 0[0;1] and on day 3 with 0[0;0] in both groups (Figure 3A and B, Clinical scores of the symptoms “fatigue”(A), and “headache”(B) during the study according to a 10 point VAS assessment by the patients). In addition to the clinical symptoms of TP, which were assessed on the basis of the patient’s complaints, the otorhinolaryngologist assessed the intensity of hyperemia of the mucous membrane of the posterior pharynx and palatine tonsils using the VAS. On the day of referral, hyperemia occurred in 100% of cases and was expressed by a median score of 9 [8;10] in both groups. There was a significant difference in hyperemia on days 3 and 7 of observation. On day 3 median severity was 0 [0;0.25] points in the BNO 1030 group *vs.* 3 [1;4] points in the sage group. On day 7, all BNO 1030 patients were considered to have a normal mucosal condition. In the sage group, slight hyperemia of the posterior pharyngeal mucosa occurred in 33.3% of cases, with a final mucosal condition median score of 0 [0;1] (Figure 3C, Clinical scores of the symptoms “hyperemia of the mucous membrane”(C) during the study according to a 10 point VAS (investigator’s assessment)).

**Figure 3:**
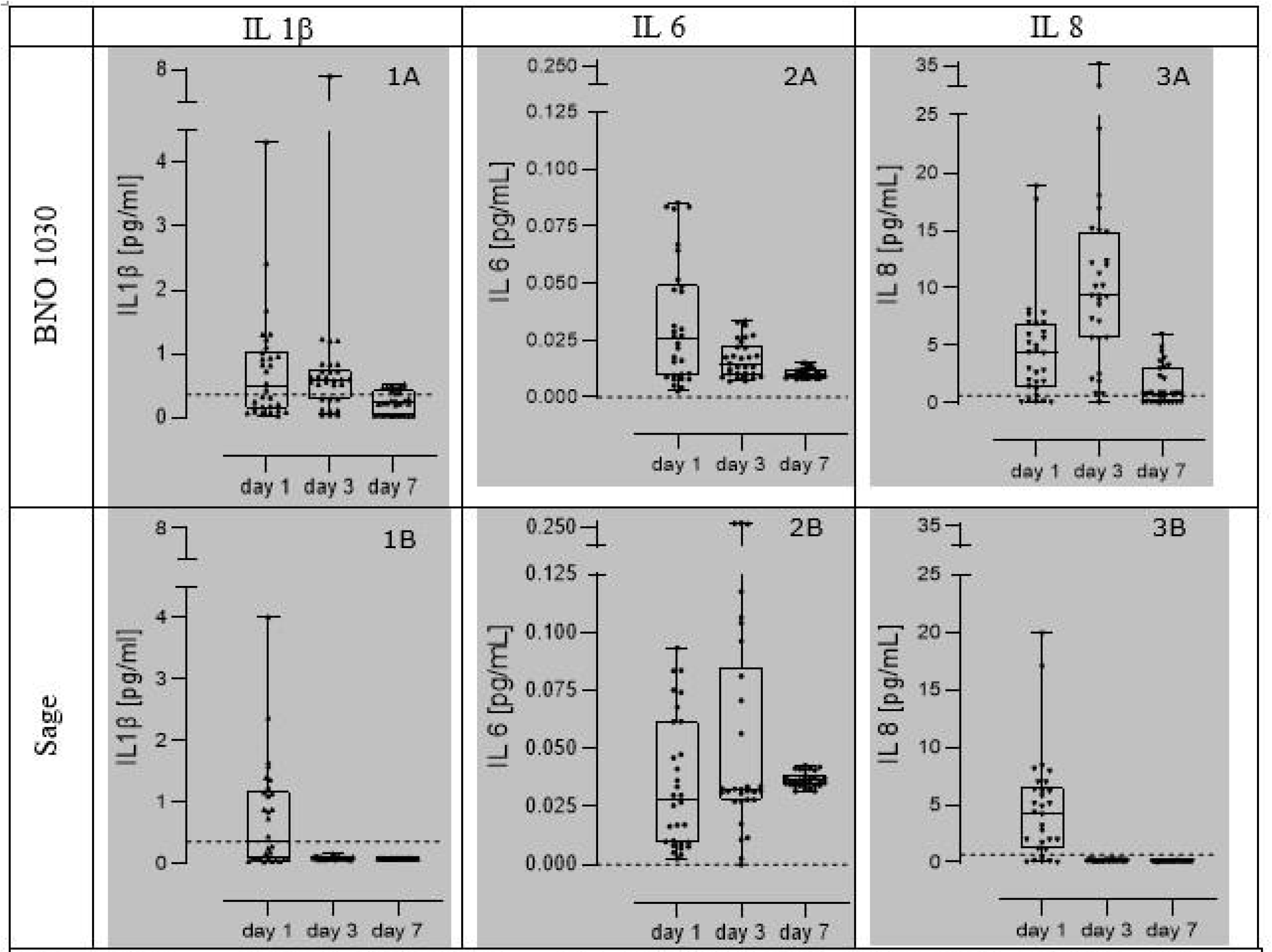
Concentrations of IL 1β (1A and B), IL 6 (2A and B), and IL 8 (3A and B) proteins from mucosal swabs of patients’ posterior pharyngeal wall and palatine tonsils after treatment with either BNO 1030 oral drops or sage lozenges on days 1,3, and 7. Box plots are median values ± 25%/75% percentiles, where whiskers represent the minimum to maximum range. Dotted lines represent the means of healthy controls. Data are presented for N=30 patients per.group.

Median body temperature was close to 37 °C on day 1 (37.1 °C and 36.8 °C) and decreased to 36.5 and 36.6 °C on day 3. On day 7 median values were nearly unaltered with median scores of 36.5 °C for BNO 1030 and 36.6 °C for sage tablets (Figure 3D, Body temperature [°C] (D) during the study). This allows to conclude that the decline value rate of the analyzed clinical manifestations of intoxication syndrome is almost identical in both groups, taking into account that the patients did not additionally take non-steroidal anti-inflammatory drugs as co-medication.

Thus, upon treatment with BNO 1030, a more rapid dynamic of intensity reduction of most of the studied symptoms of pharyngotonsillitis was observed.

### Evaluation of laboratory examination parameters

Due to absence of marked systemic inflammatory syndrome in patients with TP, all complete blood cell count parameters were within normal limits during the whole period of observation, without statistically significant difference between groups. To determine the reference values of mRNA and protein expression of IL-1β, IL-6, IL-8, IL-10, IL-17, TNF-α, lysozyme, lactoferrin and sIgA, cellular material from the mucosa of the posterior pharynx and the palatine tonsils of conditionally healthy individuals constituting the control group was taken. As a result, the level of mRNA expression of cytokines IL-10, IL-17 was below the sensitivity level of the method. Proteins of IL-6, IL-10, IL-17, TNF-α were also not detected in the supernatants of obtained cellular material by enzyme immunoassay, which indicated the absence of gene expression of these humoral factors in healthy subjects (except IL-6 mRNA) and, consequently, the absence of their proteins. The results for the remaining immunological parameters are presented in Tables 3 (Level of mRNA expression of cytokines in the oropharyngeal mucosa tissue of patients with tonsillopharyngitis in the dynamics of treatment) and 4 (Protein levels of cytokines IL-1, IL-8 lysozyme, lactoferrin, sIgA in oropharyngeal mucosa tissue of patients with tonsillopharyngitis during the treatment).

**Table 3:**
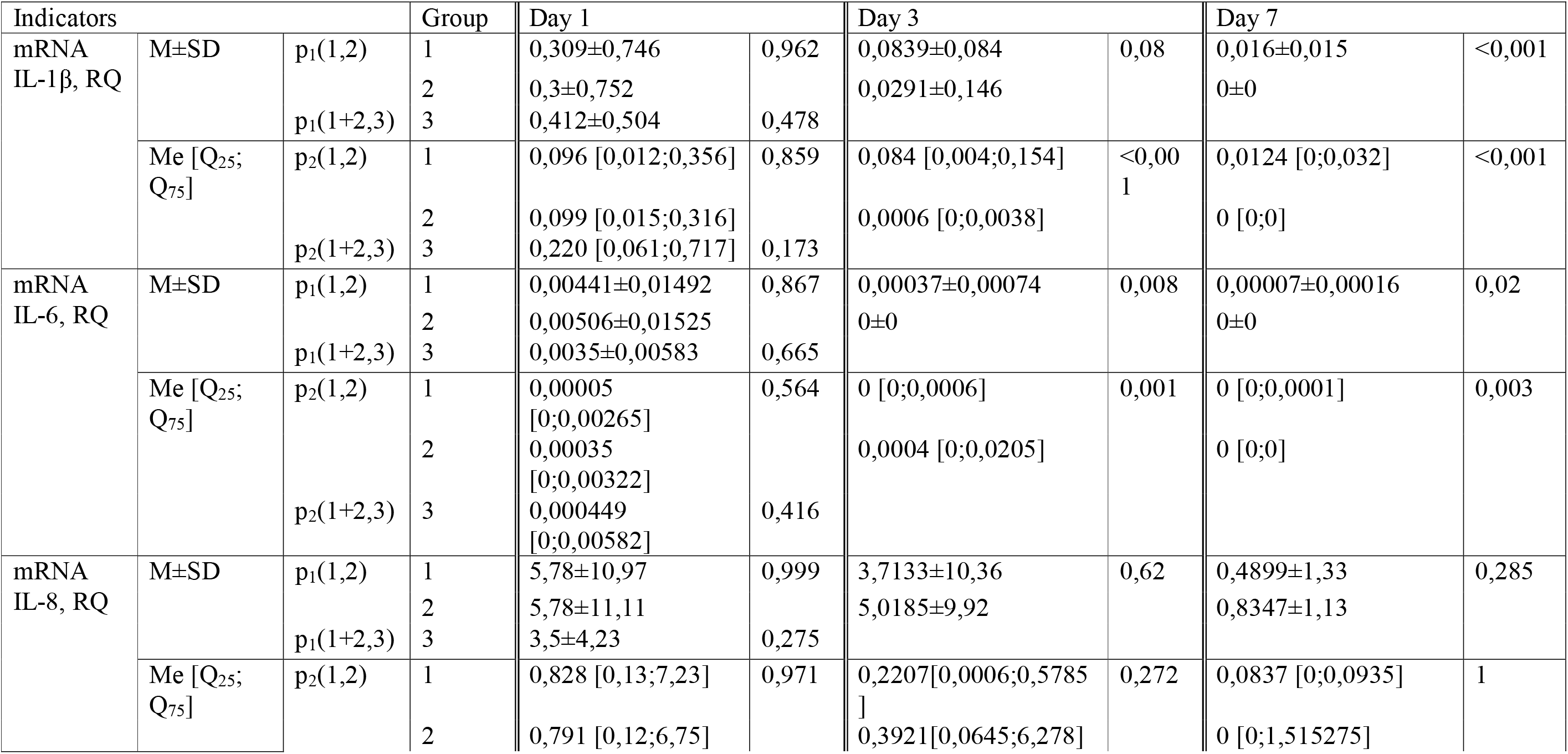
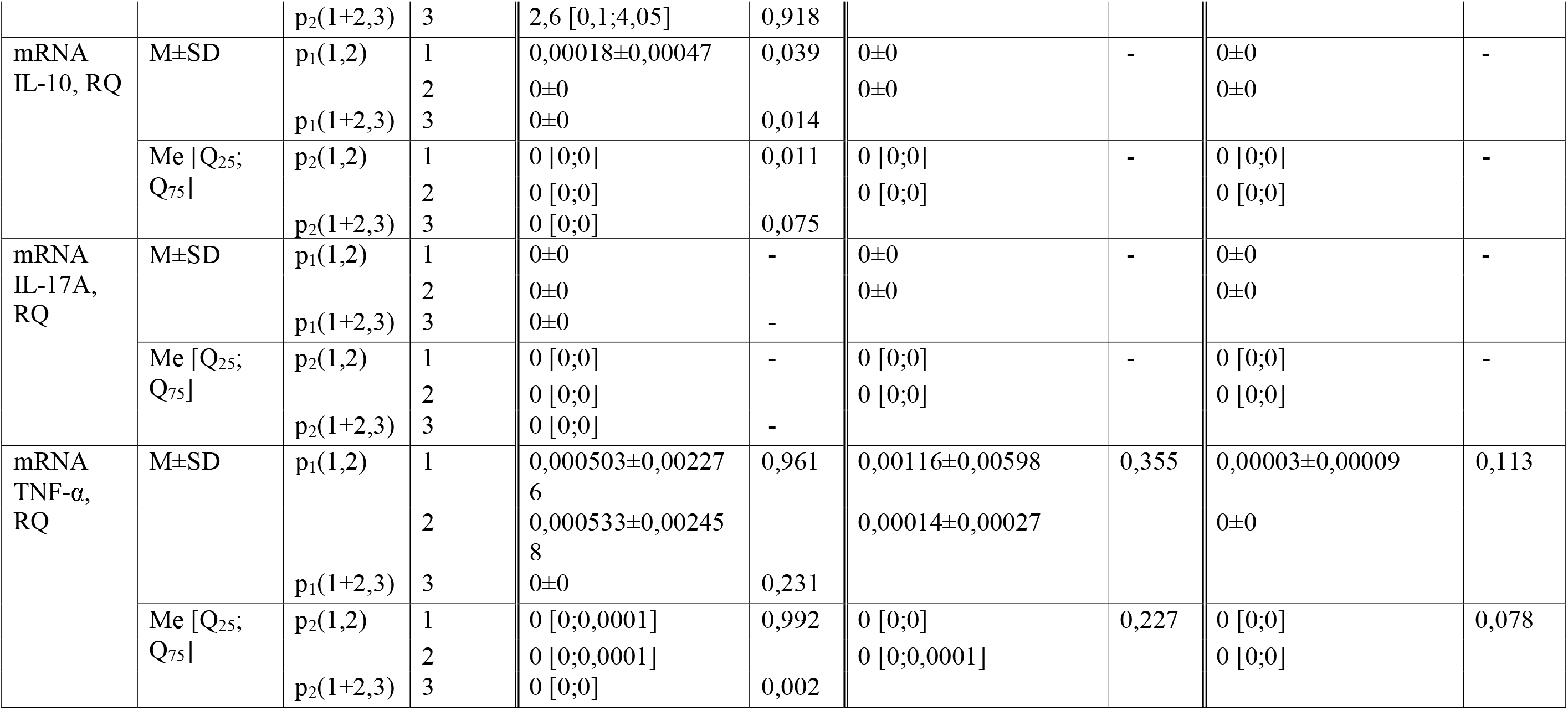
level of mRNA expression of cytokines in the oropharyngeal mucosa tissue of patients with tonsillopharyngitis in the dynamics of treatment; p1(1,2) - reliability of differences between the mean values in the first and second group, p1(1+2, 3) - significance of differences between the means in the union of the first and second groups and the third group, p2(1,2) - significance of differences between the distributions in the first and second groups, p2(1+2,3) - significance of differences between the distributions in the union of the first and second groups and the third group

The immunological parameters discussed in this section have, for the most part, a highly asymmetrical and non-normal distribution. Therefore, the median, rather than the arithmetic mean, was taken as the characteristic value, and only non-parametric methods were considered reliable when comparing data between groups.

### Dynamics of mRNA expression and protein level of IL-1**β** in patients with TP

On the first day mRNA expression and protein level of IL-1β, while there was no statistically significant difference between groups of patients with TP, were higher compared to healthy volunteers group (Tables 3 and 4). The groups differed in IL-1β protein levels from day 3 of follow up. Higher median numbers were seen in patients receiving BNO 1030 (0.784±1.283 *vs*. 0.092±0.021 or 0.594 [0.295;0.756] *vs*. 0.088 [0.081;0.096] RQ). On day 7, there was a similar difference in the comparison of the median indices (0.246±0.18 *vs*. 0.084±0.005 or 0.233 [0.037;0.436] *vs*. 0.087 [0.078;0.088] RQ) (see also Figure 4.1 A and B, Concentrations of IL 1β proteins from mucosal swabs of patients’ posterior pharyngeal wall and palatine tonsils after treatment with either BNO 1030 oral drops or sage lozenges on days 1,3, and 7). In a comparison of IL-1β mRNA expression scores, median scores of the group receiving BNO 1030 was higher than the group receiving sage on day 3 (0.0839±0.084 *vs.* 0.0291±0.146 or 0.084 [0.004;0.154] *vs*. 0.0006 [0;0.0038] RQ) and day 7 of illness (0.0155±0.0027 *vs*. 0 or 0.0124[0;0.032] *vs*. 0[0;0] RQ), where the rate was minimal. However, in both groups, the expression level of IL-1β mRNA in the oropharyngeal mucosa at the 2^nd^ and 3^rd^ visit was significantly lower than that of the control group (Figure 5.1 A and B, Relative gene expressions in comparison to control (RG) of IL 1β mRNA from mucosal swabs of patients’ posterior pharyngeal wall and palatine tonsils after treatment with either BNO 1030 oral drops or sage lozenges on days 1,3, and 7). The IL-1β protein level in the BNO 1030 group, in contrast to its mRNA expression, remained above the control group during the whole period of observation, while in group 2, starting from day 3 of observation, it was significantly below the control group.

**Table 4:** protein levels of cytokines IL-1, IL-8 lysozyme, lactoferrin, sIgA in oropharyngeal mucosa tissue of patients with tonsillopharyngitis during the treatment; p1(1,2) - reliability of differences in the mean in the first and second groups, p1(1+2. 3) - significance of differences between the means in the union of the first and second groups and the third group, p2(1,2) - significance of differences between the distributions in the first and second groups, p2(1+2,3) - significance of differences between the distributions in the union of the first and second groups and the third group

| Indicators |  |  | Group | Day 1 |  | Day 3 |  | Day 7 |  |
| --- | --- | --- | --- | --- | --- | --- | --- | --- | --- |
| IL-1β,<br>pg/mg | M±SD | p<sub>1</sub>(1,2) | 1 | 0,761±0,886 | 0,949 | 0,784±1,283 | 0,05 | 0,246±0,18 | <0,00<br>1 |
| | | | 2 | 0,746±0,886 | | 0,092±0,021 | | 0,084±0,005 | |
| | | p<sub>1</sub>(1+2,3) | 3 | 0,361±0,437 | 0,024 | | | | |
|  | Me [Q<sub>25</sub>;<br>Q<sub>75</sub>] | p<sub>2</sub>(1,2) | 1 | 0,491 [0,150;1,04] | 0,469 | 0,594 [0,295;0,756] | <0,00<br>1 | 0,233 [0,037;0,436] | 0,027 |
|  |  |  | 2 | 0,364 [0,08;1,179] |  | 0,088 [0,081;0,096] |  | 0,087 [0,078;0,088] |  |
|  |  | p<sub>2</sub>(1+2,3) | 3 | 0,188 [0;0,492] | 0,013 |  |  |  |  |
| IL-6,<br>pg/mg | M±SD | p<sub>1</sub>(1,2) | 1 | 0,0325±0,027 | 0,742 | 0,0167±0,0081 | 0,001 | 0,0105±0,0022 | <0,00<br>1 |
| | | | 2 | 0,0348±0,0282 | | 0,0653±0,0767 | | 0,0367±0,0032 | |
| | | p<sub>1</sub>(1+2,3) | 3 | 0±0 | <0,00<br>1 | | | | |
|  | Me [Q<sub>25</sub>;<br>Q<sub>75</sub>] | p<sub>2</sub>(1,2) | 1 | 0,0253<br>[0,0095;0,049] | 0,779 | 0,0141 [0,0099;0,0226] | <0,00<br>1 | 0,0097<br>[0,0086;0,0119] | <0,00<br>1 |
|  |  |  | 2 | 0,0284<br>[0,0095;0,062] |  | 0,0322 [0,028;0,0851] |  | 0,0362<br>[0,0342;0,0389] |  |
|  |  | p<sub>2</sub>(1+2,3) | 3 | 0 [0;0] | <0,00<br>1 |  |  |  |  |
| IL-8,<br>pg/mg | M±SD | p<sub>1</sub>(1,2) | 1 | 4,693297±4,53 | 0,989 | 10,8274±8,44 | <0,00<br>1 | 1,5323±1,71 | <0,00<br>1 |
| | | | 2 | 4,6768±4,64 | | 0,1596±0,05 | | 0,1163±0,02 | |
|  |  | p<sub>1</sub>(1+2,3) | 3 | 0,6788±0,2 | <0,00<br>1 |  |  |  |  |
|  | Me [Q<sub>25</sub>;<br>Q<sub>75</sub>] | p<sub>2</sub>(1,2) | 1 | 4,386[1,409; 6,939] | 0,918 | 9,323[5,67;14,894] | <0,00<br>1 | 0,842[0,126;3,061 | <0,00<br>1 |
|  |  |  | 2 | 4,291[1,172; 6,499] |  | 0,146[0,107;0,196] |  | 0,118[0,112;0,126] |  |
|  |  | p<sub>2</sub>(1+2,3) | 3 | 0,725[0,5796;0,807] | <0,00<br>1 |  |  |  |  |
| IL-10,<br>pg/mg | M±SD | p<sub>1</sub>(1,2) | 1 | 0±0 | - | 0±0 | - | 0±0 | - |
|  |  |  | 2 | 0±0 |  | 0±0 |  | 0±0 |  |
|  |  | p<sub>1</sub>(1+2,3) | 3 | 0±0 | - |  |  |  |  |
|  | Me [Q<sub>25</sub>;<br>Q<sub>75</sub>] | p<sub>2</sub>(1,2) | 1 | 0 [0;0] | - | 0 [0;0] | - | 0 [0;0] | - |
|  |  |  | 2 | 0 [0;0] |  | 0 [0;0] |  | 0 [0;0] |  |
|  |  | p<sub>2</sub>(1+2,3) | 3 | 0 [0;0] | - |  |  |  |  |
| IL-17A,<br>pg/mg | M±SD | p<sub>1</sub>(1,2) | 1 | 0±0 | - | 0±0 | - | 0±0 | - |
|  |  |  | 2 | 0±0 |  | 0±0 |  | 0±0 |  |
|  |  | p<sub>1</sub>(1+2,3) | 3 | 0±0 | - |  |  |  |  |
|  | Me [Q<sub>25</sub>;<br>Q<sub>75</sub>] | p<sub>2</sub>(1,2) | 1 | 0 [0;0] | - | 0 [0;0] | - | 0 [0;0,0001] | - |
|  |  |  | 2 | 0 [0;0] |  | 0 [0;0] |  | 0 [0;0] |  |
|  |  | p<sub>2</sub>(1+2,3) | 3 | 0 [0;0] | - |  |  |  |  |
| TNF-α,<br>pg/mg | M±SD | p<sub>1</sub>(1,2) | 1 | 0±0 | - | 0±0 | - | 0±0 | - |
|  |  |  | 2 | 0±0 |  | 0±0 |  | 0±0 |  |
|  |  | p<sub>1</sub>(1+2,3) | 3 | 0±0 | - |  |  |  |  |
|  | Me [Q<sub>25</sub>;<br>Q<sub>75</sub>] | p<sub>2</sub>(1,2) | 1 | 0 [0;0] | - | 0 [0;0] | - | 0 [0;0] | - |
|  |  |  | 2 | 0 [0;0] |  | 0 [0;0] |  | 0 [0;0] |  |
|  |  | p<sub>2</sub>(1+2,3) | 3 | 0 [0;0] | - |  |  |  |  |
| Lysozyme<br>, pg/mg | M±SD | p<sub>1</sub>(1,2) | 1 | 10,055±11,05 | 0,975 | 19,4254±14,35 | <0,00<br>1 | 12,5558±12,21 | 0,009 |
|  |  |  | 2 | 9,9648±10,94 |  | 6,6207±0,58 |  | 6,5417±0,98 |  |
|  |  | p<sub>1</sub>(1+2,3) | 3 | 2,5103±2,22 | <0,00<br>1 |  |  |  |  |
|  | Me [Q<sub>25</sub>;<br>Q<sub>75</sub>] | p<sub>2</sub>(1,2) | 1 | 5,9713[2,2607;<br>12,597] | 1 | 14,6745[7,5655;32,032<br>7] | <0,00<br>1 | 6,2154[2,7286;27,06<br>04] | 0,976 |
|  |  |  | 2 | 6,1462[2,227;11,910<br>8] |  | 6,4886[6,1476;6,9301] |  | 6,3792[5,9863;6,999<br>6] |  |
|  |  | p<sub>2</sub>(1+2,3) | 3 | 1,7176[1,2412;2,817<br>9] |  |  |  |  |  |
| Lactoferrin, pg/mg | M±SD | p<sub>1</sub>(1,2) | 1 | 23,355±21,95 | 0,999 | 63,8352±68,73 | <0,00<br>1 | 14,2279±6,22 | <0,00<br>1 |
|  |  |  | 2 | 23,364±21,79 |  | 0,4624±0,24 |  | 0,2624±0,07 |  |
|  |  | p<sub>1</sub>(1+2,3) | 3 | 4,9629±4,11 | <0,00<br>1 |  |  |  |  |
|  | Me [Q<sub>25</sub>;<br>Q<sub>75</sub>] | p<sub>2</sub>(1,2) | 1 | 21,981[0,8815;36,80<br>06] | 0,929 | 39,6059[12,5470;96,99<br>46] | <0,00<br>1 | 14,2743[8,6722;19,1<br>39] | <0,00<br>1 |
|  |  |  | 2 | 22,177[0,8902;35,69<br>45] |  | 0,3986[0,2848;0,6928] |  | 0,2470[0,2158;0,316<br>7] |  |
|  |  | p<sub>2</sub>(1+2,3) | 3 | 3,8673[1,9359;7,651<br>1]] | 0,002 |  |  |  |  |
| sIgA, pg/mg | M±SD | p<sub>1</sub>(1,2) | 1 | 1,3626±2,8 | 0,99 | 5,2367±4,51 | <0,00<br>1 | 5,5366±10,41 | 0,005 |
|  |  |  | 2 | 1,3716±2,79 |  | 0,0883±0,4 |  | 0,0002±0,0001 |  |
|  |  | p<sub>1</sub>(1+2,3) | 3 | 30,404±24,74 | <0,00<br>1 |  |  |  |  |
|  | Me [Q<sub>25</sub>;<br>Q<sub>75</sub>] | p<sub>2</sub>(1,2) | 1 | 0[0; 1,5342] | 0,947 | 3,8571[1,1888;7,7784] | <0,00<br>1 | 2,5338<br>[0,9883;5,3074] | <0,00<br>1 |
|  |  |  | 2 | 0[0; 1,6883] |  | 0[0;0,0016] |  | 0 [0;0] |  |
|  |  | p<sub>2</sub>(1+2,3) | 3 | 35,128[4,49;41,27] | <0,00<br>1 |  |  |  |  |

### Dynamics of mRNA expression and protein level of IL-6 in patients with TP

The groups of patients with TP did not differ in mRNA expression as well as protein level with respect to IL-6 level. However, there was a statistically significant difference to the control group, where its content was lower at the gene level, and IL-6 protein was not detected in healthy individuals. The groups differed in IL-6 mRNA expression from day 3 of follow-up. Patients taking BNO 1030 already had a lower IL-6 mRNA expression level than the control group on day 3 of the disease (0.00037±0.00074 or 0 [0;0.0006] *vs*. 0.0035±0.0011 or 0.0004 [0;0.0205] RQ), with a trend to decrease on day 7 of the disease to 0.00007±0.00016 or 0 [0;0.0001] RQ (BNO 1030). In the sage group, IL-6 mRNA expression was not detectable starting from the 2^nd^ visit. In contrast, the IL-6 protein content in oropharyngeal mucosa was higher in the patients treated with sage tablets on day 3 (0.0167±0.0081 *vs*. 0.0653±0.0767 or 0.0141 [0.0099;0.0226] *vs*. 0.0322 [0.028;0.0851]) and on day 7 of follow-up (0.0105±0.0022 *vs*. 0.0367±0.0032 or 0.0097 [0.0086;0.0119] *vs*. 0.0362 [0.0342;0.0389] RQ) (see also Figures 4.2A and B, Figures 5.2A and B).

### Dynamics of mRNA expression and protein level of IL-8 in patients with TP

The mRNA and protein levels of IL-8 mRNA and protein expression also differed in the patient groups with TP from those of healthy volunteers, but did not differ among themselves. IL-8 mRNA and protein were lower in the control group than in the main groups on day 1 of the disease. In contrast, IL-8 protein levels were higher in the group of patients treated with BNO 1030 and on day 3 (10.8274±8.44 *vs*. 0.1596±.05 or 9.323 [5.67;14.894] *vs*. 0.146 [0.107;0.196] RQ) and on day 7 (1.532±1.71 *vs*. 0.116±0.02) or 0.842 [0.126;3.061 *vs.* 0.118 [0.112;0.126] RQ (Figure 4 A and B, Concentrations of IL 8 proteins from mucosal swabs of patients’ posterior pharyngeal wall and palatine tonsils after treatment with either BNO 1030 oral drops or sage lozenges on days 1,3, and 7). In the BNO 1030 group, IL-8 protein amount was significantly higher than normal on day 3 of the disease, on day 7 of the disease there was a tendency for normalization, but there was still a significant difference to the control group. In the sage group the IL-8 protein level was lower than normal from day 3 of the disease.

**Figure 4:**
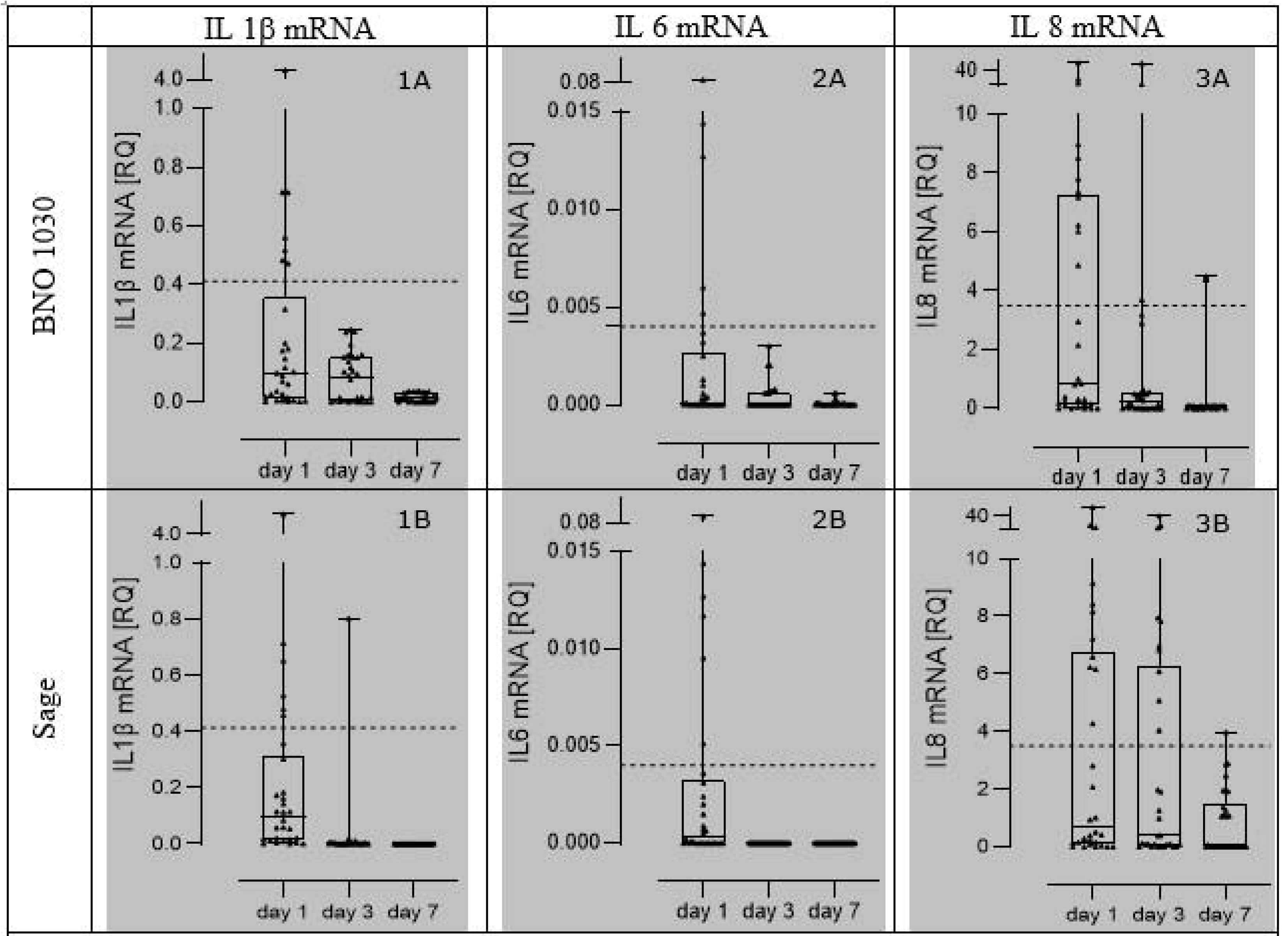
relative gene expressions in comparison to control (RG) of IL lβ mRNA (1A and B), IL 6 mRNA (2A and B), and IL 8 mRNA (3 A and B) from mucosal swabs of patients’ posterior pharyngeal wall and palatine tonsils after treatment with either BNO 1030 oral drops or sage lozenges on days 1,3, and 7. Box plots are median values ± 25%/75% percentiles, where whiskers represent the minimum to maximum range. Dotted lines represent the means of healthy controls. Data are presented for N=30 patients per group.

**Figure 5:**
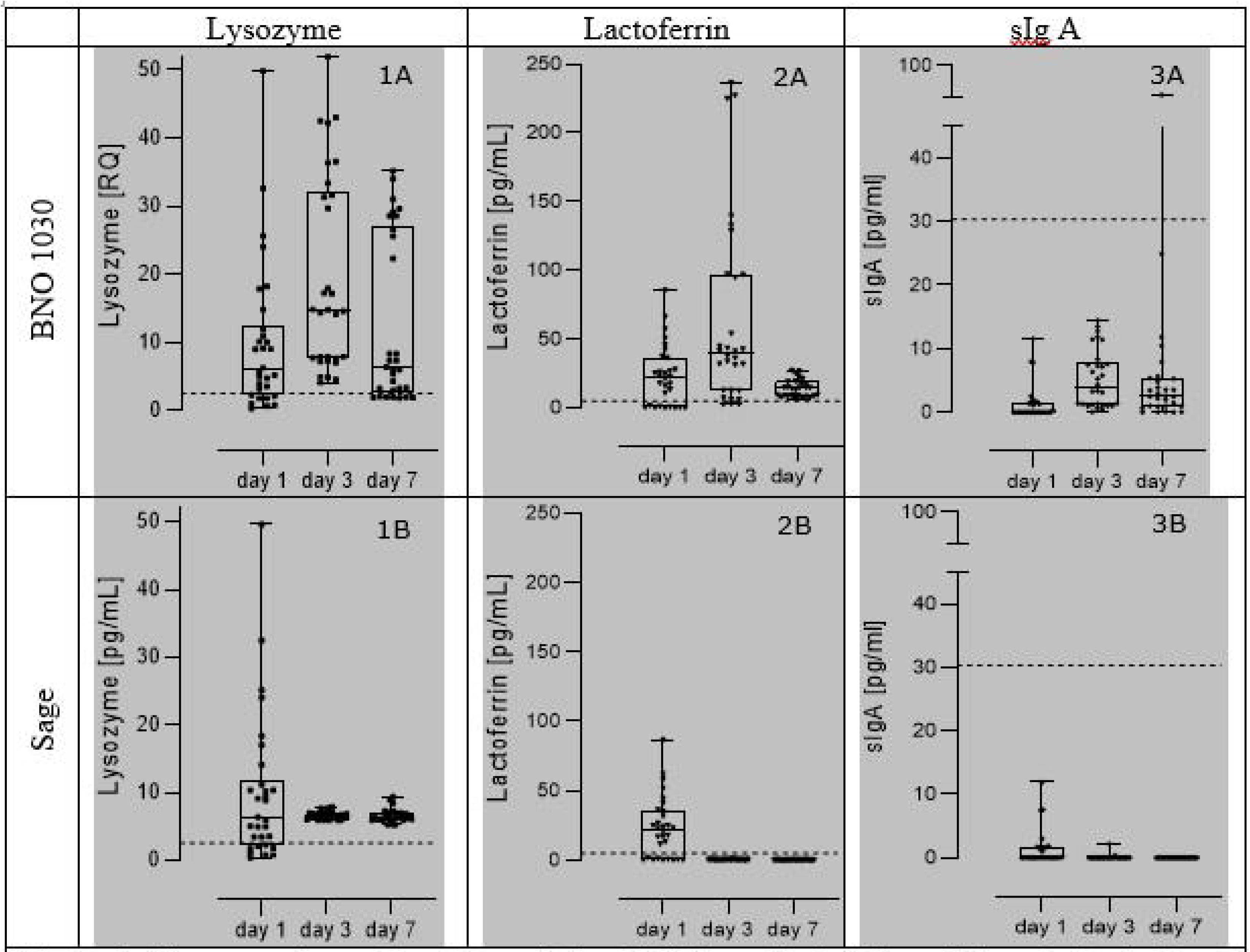
Concentrations of Lysozyme (1A and B), Lactoferrin (2A and B), and sIg A (3A and B) from mucosal swabs of patients’ posterior pharyngeal wall and palatine tonsils after treatment with either BNO 1030 oral drops or sage lozenges on days 1,3, and 7. Box plots are median values ± 25%/75% percentiles, where whiskers represent the minimum to maximum range. Dotted lines represent the means of healthy controls. Data are presented for N=30 patients per group.

Together with that, the elevated mRNA values of this cytokine persisted on day 3 of treatment, and on day 7 the IL-8 mRNA expression values were lower than those of the control group in both groups with TP.

When comparing the BNO 1030 and the sage group in comparison, the patients group 2 treated with sage therapy had significantly higher IL-8 mRNA expression levels on day 3 (3.7133±10.36 *vs*. 5.0185±9.92 or 0.2207 [0.0006;0.5785] *vs*. 0.3921[0.0645; 6.278] PQ) and on day 7 of disease (0.4899±1.33 *vs*. 0.8347±1.13 or 0.0837[0; 0.0936] *vs*. 0 [0; 1.515275] PQ) (Figure 5.3 A and B, Relative gene expressions in comparison to control (RG) of IL 8 mRNA from mucosal swabs of patients’ posterior pharyngeal wall and palatine tonsils after treatment with either BNO 1030 oral drops or sage lozenges on days 1,3, and 7).

### Dynamics of mRNA expression and protein level of TNF-**α** in patients with TP

TNF-α protein was not detected in any group, including controls, during the whole period of observation. Regarding TNF-α mRNA expression, certain values of this cytokine were obtained in groups of patients with TP, with no significant difference between groups at day 1 of the disease. No TNF-α mRNA expression was detected in healthy volunteers. On day 3 of the follow-up there were significantly higher values of TNF-α mRNA expression in the group of patients receiving BNO 1030, when comparing average values (0.00116±0,00598 *vs*. 0.00014±0,00027) RQ. On day 7 there was no significant difference between the groups (Table 3, Level of mRNA expression of cytokines in the oropharyngeal mucosa tissue of patients with tonsillopharyngitis in the dynamics of treatment).

### Dynamics of mRNA expression and protein levels of IL-10 and IL-17A in patients with tonsillopharyngitis

These interleukins yielded zero levels for both protein and mRNA expression, so no comparative analysis was performed.

### Dynamics of lysozyme levels in patients with TP

In healthy individuals of the control group the lysozyme content was significantly lower than that of patients with TP on the first day of treatment. It is remarkable, that a sharp increase of this index was observed on the background of BNO 1030 administration in comparison with the group of patients treated with sage from day 3 (19.4254±14.35 *vs*. 6.6207±0.58 or 14.6745 [7.5655;32.0327] *vs*. 6.4886 [6.1476;6.9301] pg/mL). This trend continued until day 7 (12.5558±12.21 *vs*. 6.5417±0.98 or 6.2154 [2.7286;27.0604] *vs*. 6.3792 [5.9863;6.9996] pg/mL) (Figure 6.1 A and B, Сoncentrations of Lysozyme from mucosal swabs of patients’ posterior pharyngeal wall and palatine tonsils after treatment with either BNO 1030 oral drops or sage lozenges on days 1,3, and 7). Throughout the follow-up period, lysozyme values in the TP patient groups were significantly higher than in the control group.

### Dynamics of lactoferrin levels in patients with TP

Lactoferrin innate immune response factor values in patients with TP were also significantly higher than in healthy controls on day 1 of the disease, without any significant difference between the treatment groups. In the course of treatment with BNO 1030, a prominent increase of this index on the 3^rd^ day was observed. At the same time, a sharp decrease was detected during treatment with sage (63.8352±68.73 *vs*. 0.4624±0.24 or 39.6059 [12.5470;96.9946] *vs*. 0.3986 [0.2848;0.6928]). Lactoferrin levels in the sage group were below normal ones from day 3 onwards, with decline continuing at day 7 of illness (14.2279±6.22 *vs*. 0.2624±0.07 or 14.2743 [8.6722;19.139] *vs*. 0.2470 [0.2158;0.3167] pg/ml) (Figure 6.2 A and B, Сoncentrations of Lactoferrin from mucosal swabs of patients’ posterior pharyngeal wall and palatine tonsils after treatment with either BNO 1030 oral drops or sage lozenges on days 1,3, and 7).

### Dynamics of sIgA in patients with TP

On the first day of treatment, sIgA levels in the oropharyngeal mucosa were significantly lower in patients with TP compared to healthy controls, with no significant difference between the treatment groups. There was an increase in this index with BNO 1030 and a further decrease with sage on day 3 (5.2367±4.51 *vs*. 0.0883±0.4 or 3.8571 [1.1888;7.7784] *vs*. 0 [0;0.0016] pg/mL). On day 7 of the disease, the amount of sIgA produced by the oropharyngeal mucosa in the BNO 1030 group remained at the same level as on day 3, while in the sage group this immune response factor was not detectable in most cases (86.7%) (5.5366±10.41 *vs*. 0.0002±0.0001 or 2.5338 [0.9883;5.3074] *vs*. 0[0;0]) (Figure 6.3 A and B, Сoncentrations of sIg A from mucosal swabs of patients’ posterior pharyngeal wall and palatine tonsils after treatment with either BNO 1030 oral drops or sage lozenges on days 1,3, and 7).

Please note that, **table 2** shows 40 values of statistical significance of p for pairwise comparisons, of which 14 were significant with p<0.001, 17 with p<0.01 and 20 with p<0.05. **Table 3** shows 36 values of statistical significance of p for pairwise comparisons, of which 3 were significant with p<0.001, 7 with p<0.01 and 11 with p<0.05. **Table 4** shows 47 values of statistical significance of p for pairwise comparisons, of which 26 were significant with p<0.001, 30 with p<0.01 and 33 with p<0.05. Since when using the Bonferroni correction, in order to assert the presence of statistically significant differences with p<0.05 for n comparisons, it is enough that there is at least one difference with p<0.05/n, and 0.001/40, 0.001/36 and 0.001/47 are less than 0.05 and for all three tables there were significant differences with . p<0.001. It is also possible to estimate the expected number of false positive connections. In total, 46 differences with p<0.001 were obtained from 123 comparisons. If we accept the hypothesis that there are no significant differences, then the probability that false-positive differences with p<0.001 will be more than one 1 is approximately 0.007, and the probability that there will be more than two such differences is approximately 0.0003.

## Discussion

The main task of therapy of acute inflammation in oropharynx is the reduction of duration and severity of the pathological process. It is a typical process, little dependent on the etiologic factor, in the form of local production of pro-inflammatory and then anti-inflammatory cytokines in response to the causative agent. Therefore, it is necessary to counteract the progression of the inflammatory process. It should be preferred to localize the inflammation on the level of oropharyngeal mucosa and to avoid triggering a systemic inflammatory response. The active use of herbal medicines for the treatment of acute TP can help to avoid the problem of irrational use of systemic and topical antimicrobials, which will prevent the growth of resistant strains of microorganisms and the imbalance of the normal microflora of the oropharyngeal mucosa.

In the present study, we had 2 main objectives: 1) to study the clinical efficacy of monotherapy of acute TP or exacerbation of chronic TP without a pronounced systemic inflammatory syndrome with BNO 1030; 2) to investigate anti-inflammatory properties of the preparation BNO 1030 and its effect on indices of local immunity of oropharyngeal mucosa. Please be noted that since parametric statistical methods were used for groups’ comparison, and the size of groups of 30 subjects was chosen as minimally corresponding the requirements of parametric statistics, it is necessary to be critical of the highly reliable statistical significance obtained (at the level of p<0.001), since in this case the errors in calculating the statistical significance of p at the level of several thousandths can transfer the result from significant with p<0.001 to significant with p<0.01. Taken sample size of 30 subjects is correct for searching for statistically significant differences with p= 0.05.

We used local monotherapy with sage lozenges as a comparator. There were 30 patients in each group, who did not differ in general clinical and laboratory parameters at the start of the treatment. Results of treatments in each group were compared by measuring the severity of painful swallowing, throat irritation, feeling of an oropharyngeal foreign body, dry cough, fatigue, headache, fever and hyperemia in the posterior pharynx using a 10-point VAS scale on day 1, day 3 and day 7. In conclusion we observed an regress of TP typical symptoms and relevant effects on proinflammatory cytokines and factors of the local immunity.

In the last years results of different clinical trials have been published, confirming the beneficial effects of BNO 1030 in different populations. BNO 1030 significantly decreased the symptoms of acute TP and proved a decreased need for analgetic therapy and the prescription of antibiotics [13,14]. In our study, there were also positive results for the efficacy of therapy in both groups. All patients had no significant complaints by day 7 of follow-up, none of them had progressed inflammation or developed any complications, and all patients did not require further treatment. However, a significant difference were noted in a more rapid reduction of symptoms such as painful swallowing, feeling of a foreign body in the oropharynx, throat irritation and intensity of hyperemia of the posterior pharyngeal mucosa during treatment with BNO 1030 on day 3 of follow-up compared to the patient group that received sage. On day 7 of follow-up, all analyzed clinical manifestations of TP were absent during treatment with BNO 1030, while in the sage group, symptoms of painful swallowing, feeling of foreign body in oropharynx, throat irritation, intensity of hyperemia of the mucous membrane of posterior pharynx (Me=0 [0;1] points) and dry cough (Me=0 [0;0.5] points) were still mild in some cases. Thus, treatment with BNO 1030 showed the maximum rapid reduction in duration of both catarrhal syndrome of oropharyngeal mucosa and manifestations of systemic inflammatory syndrome in comparison with treatment with sage.

The second purpose of our study was to investigate anti-inflammatory properties of the preparation BNO 1030 and its effect on indices of local immunity of oropharyngeal mucosa. A stage of the local inflammatory process course can be indicated by the analysis of proinflammatory cytokines content directly in the oropharyngeal mucosa. We determined both mRNA expression by PCR and protein expression by ELISA of cytokines IL-1β, IL-6, IL-8, IL-10, IL-17, TNF-α, in scrapings from the mucosa of the posterior pharyngeal wall and palatine tonsils of patients with TP. These characteristic factors are supported by a validation study by Geißler et al. 2020 and Savlevich et al. 2021 [11,15]. In addition, the protein levels of the most important factors of the local immunity of the lysozyme, lactoferrin and sIgA were determined by ELISA. Given the absence of reference values of all the above immunological indicators for the oropharyngeal mucosa, we took samples from 30 conditionally healthy individuals (normal) as a comparator group.

The results of the laboratory tests are summarized and shown in Table 5 (Levels of cytokines and humoral effector factors in oropharyngeal mucosa samples from patients with TP before and during therapy compared with normal). We obtained zero levels for TNF-α protein, and for both protein and IL-10 and IL-17A mRNA expression at normal and throughout follow-up in all clinical groups. This is probably due to the insufficient amount of biological material or insufficient sensitivity for the test systems used in our study.

**Table 5:**
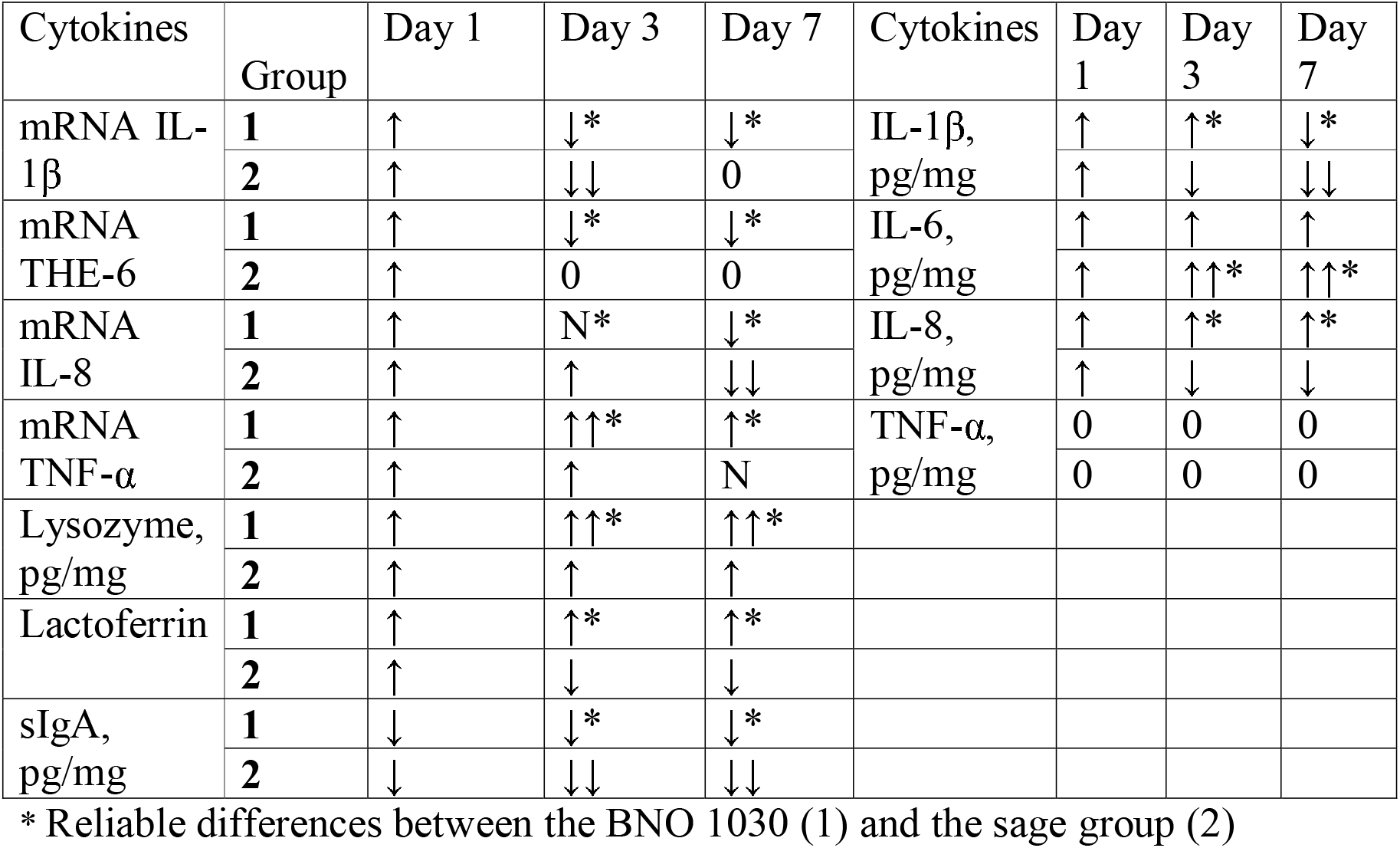
levels of cytokines and humoral effector factors in oropharyngeal mucosa samples from patients with TP before and during therapy compared with normal.

| Cytokines | Group | Day 1 | Day 3 | Day 7 | Cytokines | Day 1 | Day 3 | Day 7 |
| --- | --- | --- | --- | --- | --- | --- | --- | --- |
| mRNA IL-1β | 1 | ↑ | ↓* | ↓* | IL-1β,<br>pg/mg | ↑ | ↑* | ↓* |
|  | 2 | ↑ | ↓↓ | 0 |  | ↑ | ↓ | ↓↓ |
| mRNA THE-6 | 1 | ↑ | ↓* | ↓* | IL-6,<br>pg/mg | ↑ | ↑ | ↑ |
|  | 2 | ↑ | 0 | 0 |  | ↑ | ↑↑* | ↑↑* |
| mRNA IL-8 | 1 | ↑ | N* | ↓* | IL-8,<br>pg/mg | ↑ | ↑* | ↑* |
|  | 2 | ↑ | ↑ | ↓↓ |  | ↑ | ↓ | ↓ |
| mRNA TNF-α | 1 | ↑ | ↑↑* | ↑* | TNF-α,<br>pg/mg | 0 | 0 | 0 |
|  | 2 | ↑ | ↑ | N |  | 0 | 0 | 0 |
| Lysozyme,<br>pg/mg | 1 | ↑ | ↑↑* | ↑↑* |  |  |  |  |
|  | 2 | ↑ | ↑ | ↑ |  |  |  |  |
| Lactoferrin | 1 | ↑ | ↑* | ↑* |  |  |  |  |
|  | 2 | ↑ | ↓ | ↓ |  |  |  |  |
| sIgA,<br>pg/mg | 1 | ↓ | ↓* | ↓* |  |  |  |  |
|  | 2 | ↓ | ↓↓ | ↓↓ |  |  |  |  |

\* Reliable differences between the BNO 1030 (1) and the sage group (2)

Regarding the levels of detectable cytokines at day 1 of follow-up, there was an increase in mRNA expression and protein content of IL-1β, IL-6 and IL-8, mRNA expression of TNF-α as well as for protein of lysozyme and lactoferrin compared to healthy individuals in TP. Similar effects could be observed in studies focusing for defensins as a first line defense mechanisms on the tonsils [16,17]. The level of sIgA was low, which is consistent with our previous studies [11]. Taking into account the fact, that on day 1 of the observation patients are in the acute phase of TP and didn’t receive any therapy, the above mentioned changes can be considered as those characterizing the state of local immunity on the background of acute infectious process in oropharyngeal mucosa.

These changes are to be expected and have been previously described for acute respiratory viral infections [5]. Indeed, all 4 cytokines (IL-1β, IL-6, IL-8, TNF-α) belong to the group of inducible pro-inflammatory factors, although their roles in inflammation differ. IL-1β has many biological effects (pleiotropic action), but at the onset of inflammation it is responsible for the activation of innate immune cells at the point of injury, and at the systemic level for fever. IL-6 and TNF-α are also pleiotropic cytokines; in inflammation, they regulate the interaction between cells of the immune system for the proper development of the immune response cascade. The main task of IL-8 in inflammation is to regulate the directed migration of inflammatory cells (neutrophils and macrophages) to the site of injury (chemotaxis) to eliminate damaging agents. In addition, IL-8 regulates the release of lysozyme, lactoferrin and other antimicrobial factors from neutrophil granules [18].

Lysozyme and lactoferrin are factors of innate immunity with direct action. They are not specific to a particular pathogen. Lysozyme is an enzyme that degrades peptidoglycan, a major component of most cell walls of Gram-positive and Gram-negative bacteria. The action of lysozyme disrupts the integrity of the bacterial cell wall and results in bacterial death. In turn, there is an activation of pattern recognition receptors, including nucleotide-binding oligomerization domain-like (NOD)1 and NOD2 receptors, Toll-like receptors (TLRs), release of pathogen-associated molecular patterns (PAMPs) that leads to phagocyte recruitment being active participants of antiviral response [19]. Phagocytosis is also relevant for the regeneration of the mucosa after the inflammatory process, mediated by M2 macrophages and IL-10. At the same time, lysozyme significantly inhibits the synthesis of TNF-α and IL-6 by macrophages, providing a nonspecific anti-inflammatory effect [20].

The cationic protein lactoferrin also has a wide range of activities: antibacterial, antiviral, antifungal. Interaction with negatively charged cell surfaces prevents viral adhesion and subsequent penetration of viruses into the cell. At the same time, lactoferrin decreases intracellular reactive oxygen species and improves cellular antioxidant capacity [21]. Next to this antiviral capacities, lactoferrin is also known for its ability to activate NK cells of the antiviral response [22].

sIgA is a major effector humoral factor of adaptive immunity in barrier tissues. It provides protection of mucosal surfaces by preventing viruses and bacteria from attaching to the mucosal epithelium of the upper respiratory tract [23]. It should be considered that there is a reciprocal dependency between the amount of local pathogens and the level of sIgA. The greater the number of pathogenic agents is, the lower the sIgA level will be at the onset of the disease. As the pathological process progresses, its level should rise due to increased synthesis by mucosal B-cells. sIgA inhibit the reproduction of the pathogen at the entrance gate of the infection and prevent its escape from the body in the active form, thus preventing spreading of infection. They stimulate the subsequent attack of cytotoxic cells in case of local invasion [23]. Thus, at the onset of acute TP or exacerbation of chronic TP, as expected, a pattern of local activation of innate and adaptive immunity with an increase of regulatory (cytokines) and direct effector factors (lysozyme, lactoferrin) was observed.

On the 3rd day of the follow-up, in the group treated with BNO 1030, a decrease of IL-1β and IL-6 mRNA expression below the control group level, normalization of IL-8 mRNA level according to the arithmetic mean values and increase of TNF-α mRNA as compared to the 1^st^ day, were observed. Protein levels of IL-1β did not change significantly from day 1 and were above the control group, IL-6 protein levels were lower than on day 1 but above the control group, and there was a significant increase in protein of IL-8. Thus, we observe a certain rearrangement of pro-inflammatory cytokines, quite natural in the immune response cascade. Previously, in the hen egg model, it was also revealed that BNO 1030 significantly suppresses the synthesis of TNF-α and IL-6 by macrophages, having an anti-inflammatory effect [24].

In the sage group on the 3rd day, the expression of IL-1β mRNA was significantly lower than in the BNO 1030 group and controls. IL-6 mRNA had zero values, IL-8 mRNA level was higher than in the BNO 1030 group and controls, and TNF-α mRNA was significantly lower than in the BNO 1030 group but not as high as in controls. Regarding proteins, there was a significant decrease of IL-1β and IL-8 in the sage group on days 3 and 7 compared to day 1 and their levels were lower than those of controls and under BNO 1030 treatment. In contrast, IL-6 protein levels increased and were higher than BNO 1030 and control values. It is worth noting the high variability of IL-8 mRNA data in the groups with TP, which resulted in a difference between the arithmetic mean and the median. Given the pilot study of this model of TP and the sample of 60 individuals, the study of the changes of this cytokine mRNA expression should be further investigated. However, a significant decrease in proinflammatory cytokine levels on day 3 of the therapy against a background of infection process cannot be regarded as an entirely positive sign.

On day 7 of follow-up, a decrease in IL-1β, IL-6, IL-8 mRNA expression of proinflammatory cytokines was observed in the BNO 1030 group compared to day 3. The values of these indicators were lower than normal. TNF-α mRNA was detected in insignificant amounts, but there was a significant difference with the control group and the sage group, where its values were zero. IL-1β protein levels fell below normal only at day 7, IL-6 and IL-8 decreased compared to day 3 but were above the control values. In the sage group, no IL-1β, IL-6 or TNF-α gene expression was detected on day 7. IL-8 mRNA levels decreased compared to day 3, were lower than controls but higher than in the BNO 1030 group. Protein levels of IL-1β and IL-8 were significantly lower than those of day 3 compared to the BNO 1030 group and control. Finally, IL-6 was also, as on day 3, higher than the BNO 1030 group and control values, although significantly lower than on day 3.

As for the factors of innate immunity, in the BNO 1030 group, there was an active increase of lactoferrin and lysozyme on the 3^rd^ day of observation, contrary to the changes in the sage group, where these parameters had a significant decrease on the same day. The amount of lysozyme was higher and the amount of lactoferrin lower than in the control group. Regarding sIgA, its values increased in the BNO 1030 on day 3, but remained below control, while in the sage group they decreased even more. Thus, under therapy with BNO 1030 an increase of factors with active antiviral and antimicrobial activity was observed on the 3^rd^ day, while in contrast the administration of sage resulted in the decrease of the level of immune protective mechanisms.

On day 7 of follow-up in the BNO 1030 group, lysozyme and lactoferrin levels decreased compared to day 3, but were above control values, and sIgA slightly increased, but was lower than control. In fact, the anti-inflammatory response has subsided against the background of the absence of clinical manifestations of TP. In the sage group, lysozyme levels slightly decreased, remaining above the control group, while minimal values were determined for lactoferrin and sIgA.

As the upper respiratory mucosa is a protective barrier and the first line of innate immunity, the replication of respiratory viruses should be limited at this level. Studies show that high levels of sIgA are important to prevent infection and protect against influenza [25].

The protective effect of sIgA is due to the ability of this antibody to inhibit the adsorption of antigens with the formation of immune complexes that neutralize the biological activity of antigens and prevent microorganisms from adhering to epithelial cells and penetrating the virus into cells. It neutralizes viruses directly in epithelial cells, binds antigens in own mucous membrane plate and excrets them through epithelium into external secretions, ridding the body of locally formed immune complexes and reduces their entry into systemic circulation [23]. Therefore, the increase of sIgA, lysozyme and lactoferrin detected by under treatment with BNO 1030 suggests an additional mechanism of action of this drug on the local immune response of the oropharyngeal mucosa.

Previous studies with BNO 1030 inhibited *in vitro* respiratory syncytial virus (RSV) replication (as determined by the plaque reduction assay in Madin-Darby canine kidney (MDCK) cells). Further immunological studies for BNO 1030 revealed a percentual increase of active phagocytic cells (e.g. macrophages) from tonsils of patients with chronic tonsillitis when incubated for 1 hour with 1:50 and 1:500 dilutions of BNO 1030, suggesting activation of antibacterial or antiviral immunity. BNO1030 also increased the number of CD56+ NK-cells, important effectors of antiviral immunity. In addition, BNO 1030 significantly induced antibody-dependent cellular cytotoxicity (25 ± 10% lytic activity in cells treated with 1:500 dilution *vs.* 10 ± 3% in control cells). It also induced the release of IFN-α and IFN-γ. BNO 1030 had *in vivo* effects on the modulation of the cytolytic activity. This was shown by antigen-challenged Wistar rats. BNO 1030 significantly increased cytolytic activity of splenocytes from rats treated with 0.3 x the human equivalent dose during 5 days. In addition, the number of antigen-specific antibody producing cells in the spleen was significantly increased [26].

We observed at the onset of acute TP or exacerbation of chronic TP, as expected, a pattern of local activation of innate and acquired immunity with an increase in regulatory (cytokines) and effector (lysozyme, lactoferrin) factors. In the further course of the inflammatory process, a certain rearrangement of proinflammatory cytokines, quite natural for the cascade of immune reactions, was observed. At the same time, just at therapy by BNO 1030 on the 3^rd^ day an increase of effector humoral immunity factors was observed, while prescription by sage, on the contrary, led to the decrease of level of immune protective mechanisms.

In summary, therapy with either BNO 1030 or sage showed a clear relief of the clinical symptoms of tonsillopharyngitis during the treatment period with recovery of the mucosal pharyngeal barrier.

Reduction of the main symptoms was significantly faster under BNO 1030 monotherapy than under sage monotherapy. In particular, in the case of BNO 1030, there were clear correlations between clinical symptoms, the onset of action and the measured local immunological parameters, the mRNA and proteins of pro-inflammatory cytokines IL-1β, IL-6, and IL-8 were decreased below the level of the healthy controls during treatment. In contrast, the effector humoral immunity factors lysozyme, lactoferrin, sIgA were activated. Therefore, compared to the sage lozenges, a broader pharmacological influence on relevant immunological mechanisms may be assumed.

To sum up we can conclude that according to the clinical picture and the change in the of the local immunity parameters, treatment of tonsillopharyngitis with an BNO 1030 extract is more preferable, since, compared with sage, the drug more quickly induce regress of TP clinical symptoms and maintains a high activity of regulatory and effector factors of the local immunity of the mucous membrane of the oropharynx, thus contributing to the anti-infectious defense of the body. Thus, the BNO 1030 extract is not only a symptomatic therapeutic, but also an immunoregulatory and anti-inflammatory remedy for the most effective response of the body to inflammation.

## Data Availability

All data produced in the present work are contained in the manuscript.

## List of abbreviations

BNO 1030: Tonsilgon N: Herbal medicinal product containing marshmallow root, chamomile flowers, horsetail herb, walnut leaves, yarrow herb, oak bark, dandelion herb
CD: cluster of differentiation
ENT: ear nose throat
FGBU: Federal State Budgetary Institution
IFN: interferon
IL: interleukin
MCP: monocyte chemotactic protein
MDCK: Madin-Darby canine kidney cells
Me: median
mRNA: messenger ribonucleic acid
NK cells: natural killer cells
NOD: nucleotide-binding oligomerization domain-like receptors
PAMPs: pathogen-associated molecular patterns
PCR: polymerase chain reaction
RANTES: regulated upon activation, normal T cell expressed and secreted
RNA: ribonucleic acid
RT-PCR: real time polymerase chain reaction
RQ: relative quantity
SD: standard deviation
sIgA: secretory immunoglobuline A
SmPC: Summary of product characteristics
TLRs: Toll-like receptors
TNF: tumor necrosis factor
TP: tonsillopharyngitis
VAS: visual analogue scale

